# Using a health equity lens to measure patient experiences of care in diverse healthcare settings

**DOI:** 10.1101/2024.01.12.24301257

**Authors:** Annette J. Browne, Colleen Varcoe, Marilyn Ford-Gilboe, C. Nadine Wathen, Erin Wilson, Vicky Bungay, Nancy Perrin

## Abstract

People who are structurally disadvantaged and marginalized often report poor health care experiences due to intersecting forms of stigma and discrimination. There are many measures of patient experiences of care, however, few are designed to measure equity-oriented care. In alignment with ongoing calls to integrate actions in support of health equity, we report on the development and testing of patient experiences of care measures that explicitly use a health equity and intersectional lens. Our analysis focuses on two different equity-oriented health care scales. The first was piloted in a primary health care setting, where patients have an ongoing relationship with providers over time. The second was piloted in an emergency department, where care is provided on an episodic basis. Item Response Theory was used to develop the scales and evaluate their psychometric properties. The primary health care scale, tested with a cohort of 567 patients, showed that providing more equity-oriented health care predicted improvements in important patient self-report health outcomes over time. The episodic scale, tested in an emergency department setting with 284 patients, showed evidence of concurrent validity, based on a high correlation with quality of care. Both scales are brief, easy-to-administer self-report measures that can support organizations to monitor quality of care using an equity lens. The availability of both scales enhances the possibility of measuring equity-oriented health care in diverse contexts. Both scales can shed light on experiences of care using an intersectional lens and equity-oriented lens, providing a nuanced understanding of quality of care.

## Introduction

Greater equity in health and health care is associated with better population health (1, 2). Achieving these aims requires focusing on those who have the least access to the social determinants of health and face the greatest barriers to health care, including those who are most structurally disadvantaged and marginalized in our societies. The idea of structural disadvantage recognizes that inequities are structural because they are embedded in social, economic, and healthcare policies and practices, and contribute to tangible, negative impacts on health, quality of life and well-being. Research shows that people who are structurally disadvantaged and marginalized experience poorer outcomes on many measures of health, and report poorer health care experiences (3–5). There is also a clear body of evidence showing that people’s health care experiences influence their timely access to healthcare, and their overall health. Importantly, a growing body of research confirms that people who have negative health care experiences or anticipate such experiences, including experiences arising from stigma and discrimination, are deterred from accessing care (3, 6–8). Therefore, measuring patient experiences of care (PEOC) and using such data to improve care are crucial to promoting equity. In a program of research and knowledge mobilization known as *<u>EQUIP Healthcare</u>*, we have been developing and testing ways to measure patient experiences of equity-oriented health care (EOHC). Grounded in a critical theoretical conceptualization of health equity, the notion of EOHC explicitly aims to: (a) address the frequent mismatches between usual approaches to care, and the needs of people most impacted by health and social inequities, (b) mitigate the impact of multiple, intersecting forms of discrimination, racism and stigma, and (c) take into account the health effects of social and structural inequities (9).

Limited attention has been given to defining what constitutes EOHC from the perspective of patients, or to developing appropriate and valid ways of measuring such care. A search of the literature shows that there are few options with regard to equity-nuanced scales to measure quality of care from the perspective of patients. Scales that attempt to measure PEOC are often limited to comparing care, or outcomes of care, for specific groups of people using a single variable, typically ethnicity, or in some jurisdictions, “race”. For example, current measures of patient-centered care, patient outcomes and experiences of care are mainly assessed through the lens of a single variable (e.g., satisfaction on a 10-point scale, or access to care with yes/no response options), and are often focused on particular ethnocultural or racialized groups (10, 11). Scales or items developed to measure experiences of care that strive to use an equity lens often focus primarily on racialized groups (e.g., in the USA, categories such as Black, Hispanic, etc.), and on groups experiencing financial strain (12, 13). Measures based on variables defined by population groups can be helpful to gauge how particular groups of people experience care. However, experiences of care are bound up with numerous factors such as feeling that one has been treated in a respectful manner (particularly for people who have had past negative experiences), and that care has been tailored to meet their particular needs and priorities (9, 14, 15). In this context, a broader approach is needed to measure EOHC.

While the limited evidence suggests that measuring equity in care settings is feasible through patient experience surveys (16), there are few options for measuring PEOC that use an intersectional perspective (8). Intersectionality offers a perspective for understanding how multiple forms of social inequity interact, and interrelate to produce relative advantage or disadvantage (17, 18). We have used an intersectional lens to highlight how inequities are experienced, typically, on the basis of inter-related, co-constituted factors and conditions -- and not solely on the basis of any one particular category, variable or group affiliation. For example, patients may perceive (or even anticipate) being inadequately cared for on the basis of being stigmatized as “drug using”, or because they are presumed to be overusing the system (i.e., if they have sought help multiples times in a short period), or for myriad intersecting reasons. In the Canadian context, Indigenous peoples are often subject to stigma related to substance use, regardless of whether or not they use substances; research shows, for example, that Indigenous people who come to emergency departments (EDs) and present with symptoms such as unsteady gait or slurred speech are often assumed to be using alcohol or other substances, and treated as such, when they may be experiencing a stroke or other serious health issues (3, 4, 19, 20). In many cases, people avoid seeking care, or leave without being seen, because of fear of being negatively judged or treated in a dismissive manner (4, 21–23). As we have shown in previous research, people who anticipate poor treatment prepare for health care encounters as carefully as they can and engage with vigilance and distrust, undoubtedly shaping those encounters (4, 11, 24). The aim in efforts to measure PEOC through an intersectional equity lens is not to determine the veracity of such experiences. Rather, the aim is to illuminate how indicators of PEOC tend to be experienced as a constellation of inseparable and intersecting experiences that are tied to issues of power and structural conditions not amenable to being measured solely on the basis of any one variable or characteristic (such as ethnicity). The purpose of this paper is to report on research focused on developing and testing measures of PEOC that explicitly uses a health equity and intersectional lens. Our analysis focuses on two different equity-oriented health care scales. The first was piloted in a primary health care (PHC) setting, where patients have an ongoing relationship with providers over time. The second was piloted in an ED, where care is provided on an episodic basis.

## Background

The program of research and knowledge mobilization known as *<u>EQUIP Health Care</u>* provided a foundation for developing and testing the effectiveness of EOHC interventions by first studying how care in the PHC sector was effectively provided to structurally disadvantaged and marginalized populations (25, 26). The process included identifying the key dimensions of EOHC, strategies to guide organizations in implementing those key dimensions (25, 27), and identification of indicators of such care relevant in PHC settings (28, 29). Guided by a framework articulating the key dimensions of EOHC and 10 strategies to support implementation, we then developed an organizational-level, multi-component health equity intervention referred to as *EQUIP Primary Health Care* [*PHC*], and tested it in four Canadian PHC settings (9, 14, 30). Building on the insights from *EQUIP PHC*, we subsequently tailored and modified the intervention (referred to as to *EQUIP Emergenc*y) to test it in three Canadian EDs (8, 31). As evidence-based and theoretically informed interventions, *EQUIP PHC* and *EQUIP Emergency* are designed to enhance organizational capacity to provide EOHC, particularly for people who experience significant health and social inequities. Throughout we have drawn on intersectionality, which emerged from Black feminist scholarship (8, 17, 32, 33), and complexity theory to draw attention to health care organizations as complex adaptive systems whose care processes can be tailored and strategically redirected to meeting the needs of people in varied contexts (34–36). As the EQUIP program of research evolves, we are continually refining our understanding of key dimensions of EOHC, which provide the basis for the EQUIP interventions. For the purposes of *EQUIP PHC* and *EQUIP Emergency*, these key dimensions were defined as including: (i) trauma- and violence-informed care (TVIC): recognizing and limiting the effects of trauma and violence, including structural violence, on peoples’ lives and care experiences; (ii) culturally-safe care approaches: reducing power imbalances, systemic racism, and discrimination; and (iii) harm reduction: preventing harms from substance use stigma, and in the process, promoting opportunities for well-being in the context of substance use (9). In our subsequent research, our team is explicitly naming our stance toward cultural safety as “antiracism” and integrating the concept of “Substance Use Health” as a non-stigmatizing approach that encompasses harm reduction. In the context of the *EQUIP Health Care* research program, Substance Use Health is used as a lens that frames substance use in relation to a spectrum that encompasses non-use, beneficial uses, occasional risks or harms, use that has ongoing or understood harms and consequences, and substance use disorders (37). It encompasses harm reduction to promote health in relation to using substances. Substance Use Health is increasingly being integrated as an essential component of health equity actions within organizations and at the point of care (37, 38).

PHC settings and EDs are critical contexts within which issues of equity and inequities must be addressed, particularly in light of ongoing reductions in community-level primary care services in most jurisdictions in Canada, with concomitant and increasing pressures on EDs to bridge the gaps in care (4, 8, 21, 39). The literature confirms that people who experience significant health and social inequities face the greatest challenges accessing primary care; consequently, people are increasingly accessing care in EDs for needs that, if resources were available, could be addressed in the PHC sector (3–5, 20, 40–42). For example, in previously published data from *EQUIP Emergency*, we showed that structurally disadvantaged groups of people were significantly less likely to have regular primary care access, and significantly more likely to have repeat ED visits, to present to EDs with health issues that were rated as lower acuity, and to present with chronic health problems (8). Additionally, as discussed by other researchers, when patients who are structurally disadvantaged (e.g., because they are precariously housed and/or living on or near the street, and/or have significant substance use issues, or have major mental health issues) seek care at the ED, the chances of experiencing negative judgements or stigmatization are high (43–45). Similarly, data from *EQUIP Emergency* showed that structurally disadvantaged groups of people reported significantly more discrimination in EDs, and rated their care more poorly than other groups (8).

Throughout the EQUIP program of research, our intention has been to invite people to describe their experiences of care in ways that assess EOHC. In our *EQUIP PHC* research, we mobilized data to develop the Equity-Oriented Health Care Scale (abbreviated to EHoCS) (14). The EHoCS was developed in primary care contexts, in which there is an assumption of an ongoing relationship between patients and the healthcare setting. Subsequently, for the *EQUIP Emergency* study we modified the scale to capture experiences of care in a single visit. Since care is provided on an episodic basis, this scale is called the Experiences of Equity during Episodic Health Care Scale (abbreviated to EEE-HC Scale). In this paper we first describe development of the original scale development for use in the primary care sector. We then discuss its ongoing adaptation for use in episodic care contexts such as EDs. We conclude with a discussion of the implications for measuring patient experiences of EOHC from an intersectional perspective, particularly in settings where patients have episodic contact across the continuum of care.

### Scale development: Equity-Oriented Health Care Scale (EHoCS) in *EQUIP Primary Health Care*

Drawing on our evolving conceptualization of the key dimensions of EOHC (9, 30), in the *EQUIP PHC* study, we used conventional scale development approaches combined with item response theory (IRT) to develop and evaluate the psychometric properties of a brief patient self-report measure of EOHC, called the EHoCS, for use in PHC and other settings where patients have ongoing contact over time. The EHoCS taps into aspects of EOHC that can be assessed using patients’ self-reports. As noted above, refinements to the key dimensions of EOHC integrating findings from our ongoing research are published in Browne et al. (2018) and further refinements are underway; however, the conceptual grounding of the EHoCS remains unchanged.

#### Item generation, pilot testing and item mapping

An initial pool of 52 items was developed to reflect domains of EOHC that align with the key dimensions, and would be amenable to patient self-report, drawing on existing research and measurement tools in the area of patient experiences of care and quality of care (28–30, 46). The five domains identified included: (1) create a welcoming, comfortable milieu (WCM); (2) promote accessibility and reduce barriers (ARB); (3) tailor care to individual context, history and experience (TIC); (4) promote emotional safety and trust (EST); and (5) convey a non-discriminatory posture (NDP). Two core/anchor items were identified for each domain. For each item, patients were asked to rate how often in the previous 12 months their PHC providers had engaged in an action reflecting EOHC, on a 5-point scale ranging from “never” to “always.” Cognitive interviews were conducted with 5 patients in one PHC setting to assess the clarity and meaning of each item from the patient’s perspective and its adequacy for measuring EOHC using patient self-reports. Based on this process and team analysis, 32 items, organized in 5 domains, were retained for psychometric testing.

In *EQUIP PHC*, we tested the EHoCS items with a cohort of patients accessing care in four PHC clinics in two Canadian provinces over two years (14). As discussed in detail in prior publications, the cohort included a diverse set of 567 patients from four different PHC clinics mandated to serve people experiencing major structural disadvantages and marginalization (due to income, geography, education, racism, ableism, and other forms of stigma and discrimination) (9, 14, 47, 48). The sample was recruited from patients who had an existing connection with the clinics. We invited patients to comment on their overall experiences of care involving all staff, versus their impressions of any one particular staff member, realizing that primary care settings are oriented to providing team-based care.

#### Psychometric evaluation of EHoCS

Structural validity of the scale was evaluated using Confirmatory Factor Analysis (CFA) in MPLUS (49) to examine the extent to which the items identified within a domain fit with the underlying construct using Chi Square and 3 fit indices: the Comparative Fit Index (CFI), Tucker Lewis Index (TLI) and Root Mean Squared Error of Approximation (RMSEA). Both CFI and TLI are incremental fit indices that compare a model with a baseline model (i.e. one with the worst fit); values range from 0 to 1, with a good fit indicated by values ≥.95 (50), with >.90 acceptable for TLI (51). RMSEA is an absolute fit index, where a value of 0 equates to an exact fit; values of < .05 are considered *close fit;* between .05 and .08 a *fair fit;* between .08 and .10 is *mediocre fit;* and > .10 a *poor fit.* Fit and modification indices were inspected to determine whether the model fit could be improved.

With 32 items in the model arranged in 5 domains, the Chi-square test for the overall model was significant (see Table 1). Given our commitment to retain those domains around which the scale is organized, we then ran separate CFAs using the items in each domain to better understand how they were contributing to the latent construct, and to potentially identify items that could be deleted. For each scale, using the modification indices, and considering theory and redundancy between items, we identified 8 items for deletion (at least one from each domain), resulting in 24 items remaining. Results of a new CFA conducted with these 24 items (organized into the same 5 domains) revealed substantially improved model fit and supported a good fit between the 5-domain structure of the scale and the item pool. Within each domain, item factor loadings ranged from .47 to .91. Thus, scores for each domain and for the overall scale were computed by summing applicable items and dividing by the number of items on the scale (range 0-4), where higher scores reflect more positive perceptions of EOHC.

**Table 1.**
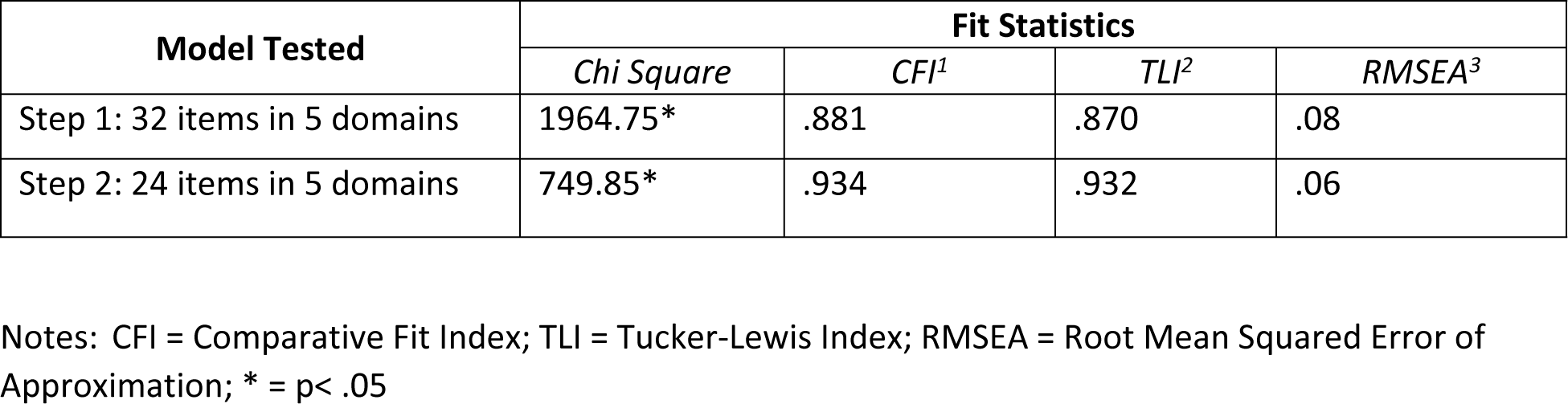
Fit indices for CFA of the EHoCS.

Internal consistency was .92 for the overall scale and .65 to .82 for each domain. However, three of the 5 domains (ARB, EST, TIC) overlapped more than expected (correlations .92-.96), suggesting that redundancy remained. For practical uses, the scale was also still quite long (i.e. 24 items). Further, the distribution of scores was highly skewed with limited variability. Ceiling effects are commonly observed in patient self-report measures focused on satisfaction with care or quality of care (52). Ceiling effects occur because many patients rate providers at the top levels possibly because of respect or social desirability, and are problematic because they tend to disguise important differences in experiences of care, including when care is sub-optimal and in need of improvement. Thus, while these analyses supported the validity and reliability of the EHoCS based on conventional psychometric testing, we recognized that a different approach was needed to further simplify the scale and improve its ability to capture variation in perceptions of EOHC in order to be useful in research, quality improvement, or decision-making contexts.

#### Improving scale precision and discrimination using IRT

To address these issues, we drew on IRT to further reduce the length of the scale while improving its ability to discriminate between different levels of EOHC (from lowest/least ideal to highest/most ideal). As an alternative to traditional psychometric testing, IRT is ideally suited to address issues of redundancy, precision and discrimination. IRT begins from an assumption that items measure a single domain, but are of varying levels of difficulty (rather than assuming similar difficulty of items as is the case in classical measurement theory) (53). Here, low difficulty items would be those that providers do very frequently, while high difficulty items are those behaviors that providers are less likely to do.

Using the 24 items retained after the CFA, we used an iterative process to compare the item characteristic curves generated for each item and IRT parameters in order to select a brief pool of items that would reflect the range of difficulty across each of the 5 domains (analyses were conducted in STATA 16.0). In making decisions about which items to retain or delete, we privileged the 2 items from each scale that had been identified as core items. Results of the IRT analysis for the final 12 items in the scale are shown in Table 2, with items organized in descending order from least to most difficult. This item pool includes 9 out of 10 core items and 3 additional items, with ten positively-worded and 2 negatively-worded items. The final scale is shown in Table 2.

**Table 2.** Item difficulty and rates of item endorsement by participants in IRT analysis.

| In the past 12 months, how often did the health care providers here*: | Difficulty | % Endorsed |
| --- | --- | --- |
| How often have you felt discriminated against by staff here, including health care providers, receptionists and others? | -1.71 | 88% |
| Have a negative attitude toward people using services because of mental health concerns? | -1.61 | 84% |
| How often did staff here treat you with courtesy and respect? | -1.12 | 79% |
| Try to make you feel as comfortable as possible? | -1.03 | 81% |
| How often did the staff here welcome you when you came for care? | -0.95 | 73% |
| Encourage you to come and see them or call when you need to? | -0.71 | 71% |
| **Seem open to talking about sensitive issues, for example, grief, mental health problems, substance use, or abuse experiences?** | -0.45 | 63% |
| **Help you to work on any barriers you have accessing health care (e.g., costs of medication or services, problems with transportation or childcare, problems getting a referral, etc.)?** | -0.14 | 54% |
| **Give you health advice that is suitable for your everyday life?** | -0.03 | 51% |
| **Try to help you to get services that are not offered here (such as social assistance, disability benefits, housing, or parenting support)?** | 0.21 | 44% |
| **Ask you about who is important in your life?** | 0.69 | 28% |
| **Ask about basic resources that affect your health, such as food, clothing, or shelter?** | 0.76 | 27% |

\*Instructions: These questions ask about your experiences with staff at this service site in the past 12 months. By staff, we mean anyone who works here including health care providers, reception staff, and others.

#### Format and scoring of the 12-item EHoCS

The EHoCS is comprised of 12 items that reflect 5 domains of EOHC. Respondents are asked to rate, in the previous 12 months, the extent to which their interactions with health care staff were equity-oriented on a 5-point scale ranging from never (0) to always (4). The EHoCS total score is a count of the number of items rated by patients as “always” occurring (for 10 positively worded items) and “never” occurring (for 2 negatively worded items), with a range of 0 to 12. Scores on the EHoCS provide an index of the degree or level of EOHC, from lower to higher. Total scores correlated with measures of overall quality of care (r= .602) and fit of care with needs (r = .599), providing evidence of concurrent validity.

In the *EQUIP PHC* study, as previously published (14), patients completed structured interviews, which included the EHoCS and other self-reported health outcome and quality of life scales, at 4 time points (baseline, 12, 18, and 24 months later). As discussed in the prior publication, using path analysis techniques, analysis with the EHoCS showed that providing more EOHC predicted improvements in important patient health outcomes 18 months later, supporting predictive validity of the EHoCs. These findings suggest that the EHoCS may be useful in measuring the possible benefits of interventions to enhance EOHC in PHC settings or in monitoring care delivery as part of Continuous Quality Improvement (CQI).

### Scale adaption and development of the Experiences of Equity during Episodic Health Care scale in *EQUIP Emergency*

The construct of EOHC underpinning *EQUIP PHC*, and *EQUIP Emergency*, and the theoretical approach underlying these studies were consistent (21, 31). Thus, we considered that many of the items in the EHoCs would be relevant to measuring patient’s self-reported experiences of care. However, given the different relationship that patients have in relation to accessing care in two different healthcare contexts -- PHC and EDs -- the response options and time frame needed to be adapted to reflect their experiences during a single, episodic visit.

#### Item generation, testing and mapping

Using the EHoCS developed for the PHC context, our research team members, who had worked with the theoretical underpinnings of EOHC for decades (including those with expertise in emergency care) adapted the items to suit the episodic care setting. Each EHoCS item was reviewed through the lens of emergency and episodic care. The team also worked with clinical practice leads at two ED sites to confirm whether the items would work in an episodic context. Two items eliminated from the EHoCS in the development of the EEE-HC Scale were “ask about basic resources that affect your health” and “have a negative attitude toward people using services because of mental health concerns”. The former was judged to be beyond the scope of usual episodic and ED practice, and the latter was too specific, with discrimination in general being a broader issue, and captured by item #4. A comparison of the EHoCS and EEE-HC items shows how, for the episodic context, we broadened from primary care specific issues. We also simplified and streamlined the questions. A review by diverse stakeholders suggested that using a 5-point scale would require a level of discernment not easily made during an episodic, and often brief, health care encounter; consequently, we changed the response option to a simple yes/no. Ten adapted items were used with the binary response option for our initial testing in *EQUIP Emergency*, as shown in Table 3.

**Table 3.** Original Experiences of Equity during Episodic Health Care (EEE-HC) Scale items.

| During this visit, did staff: | Yes | No |
| --- | --- | --- |
| 1. make you feel welcome? | ☐ | ☐ |
| 2. try to make you as comfortable as possible? | ☐ | ☐ |
| 3. treat you with courtesy and respect? | ☐ | ☐ |
| 4. discriminate against you? | ☐ | ☐ |
| 5. seem open to talking about what is important to you? | ☐ | ☐ |
| 6. learn enough about you to give useful advice? | ☐ | ☐ |
| 7. give you advice that is suitable for you? | ☐ | ☐ |
| 8. learn about problems you might have getting services (e.g., costs, transportation, getting a referral, etc.)? | ☐ | ☐ |
| 9. try to help you get services you need? | ☐ | ☐ |
| 10. encourage you to return if you need to? | ☐ | ☐ |

In the context of *EQUIP Emergency*, and embedded in the larger patient survey, we tested these 10 items at one of our three hospital sites, Surrey Memorial Hospital (SMH), during a wave of patient surveys (8, 21, 31). Recruitment for the larger *EQUIP Emergency* study began on November 28, 2017 and ended on November 12, 2020. Informed consent was documented on signed consent forms for all participants.

SMH is the largest ED in the Canadian province of British Columbia, and serves diverse suburban communities, including high proportions of newcomers and refugees, with many who speak a language other than Canada’s two official languages, English or French, at home (54, 55). In Canada, “newcomers” is used as a preferred term to indicate people who were not born in Canada; this includes people classified by the Canadian federal government as immigrants or refugees (56). The hospital also serves a large urban Indigenous population of 16,300 people, including those who self-identify as First Nations, Métis or Inuit (57). Due to COVID-19 pandemic restrictions and mandated requirements to halt data collection, it was not possible to administer the EEE-HC Scale at the two other ED sites involved in the *EQUIP Emergency* study.

Surveys were conducted with 284 patients, during which they were asked about their experiences of receiving care during their visit to the ED. Research Assistants, trained in equity-oriented approaches including strategies for working respectfully with people who experience significant inequities (and are thus often not included in research), conducted the surveys and gathered patients’ feedback on the clarity, meaning and response options to the items. The recruitment efforts resulted in a sample that was diverse, and was generally representative of the populations served by SMH. This included representation from people over 65, Indigenous people, people experiencing precarious housing, people born outside of Canada, and people who found it difficult to live on their income (Table 4). The entire sample is described more fully elsewhere (8).

**Table 4.**
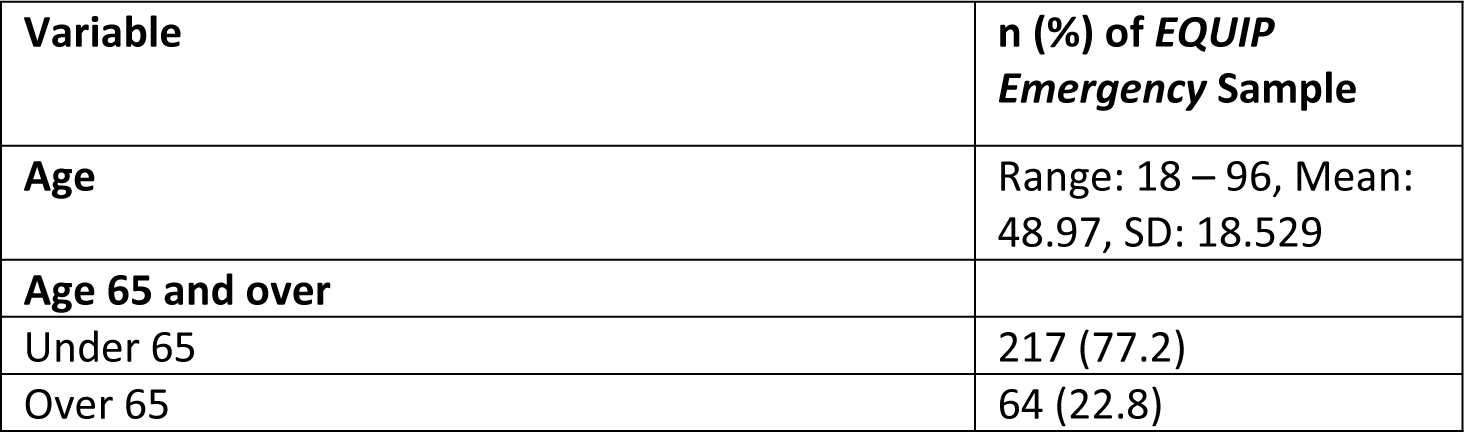
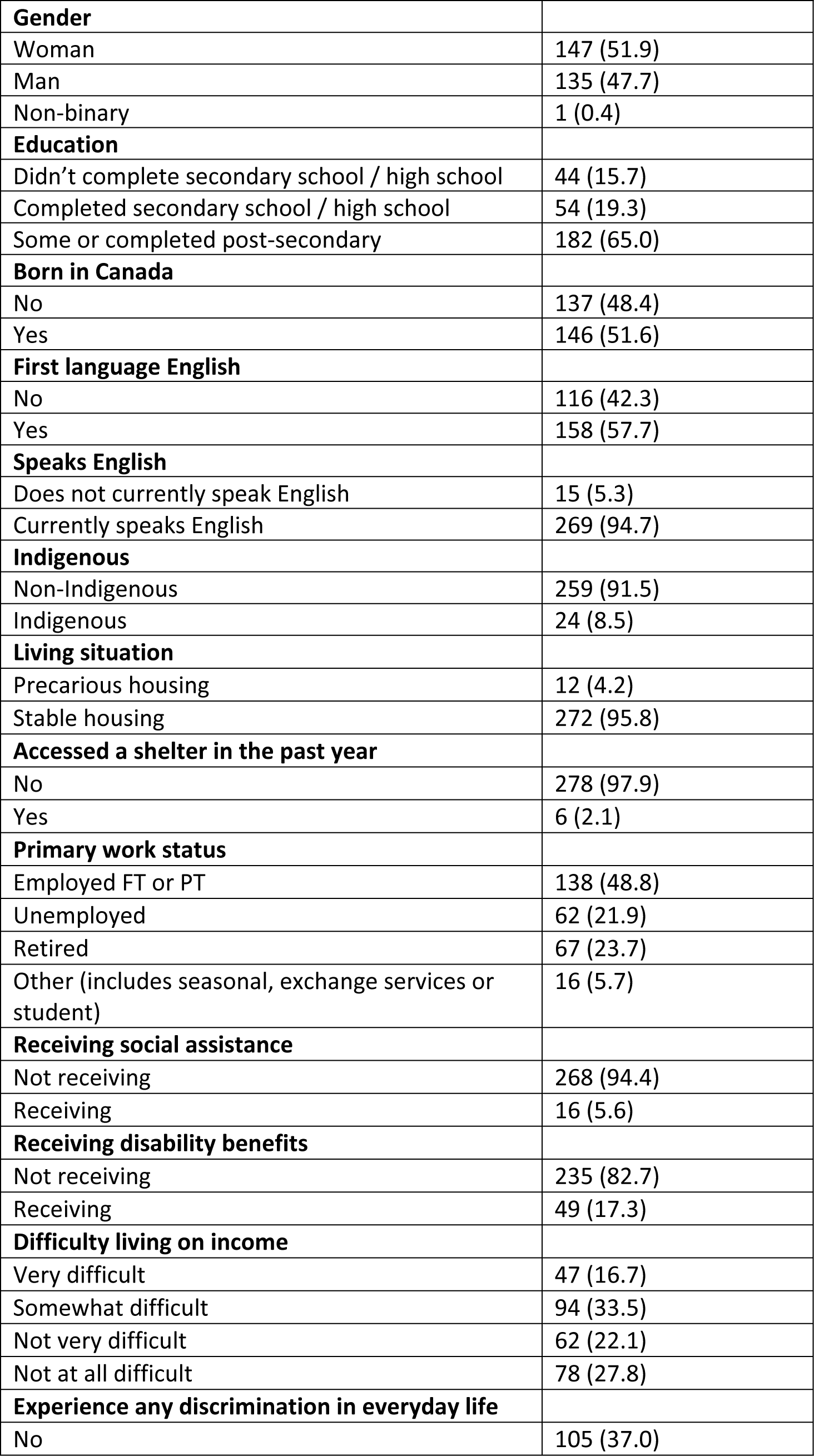
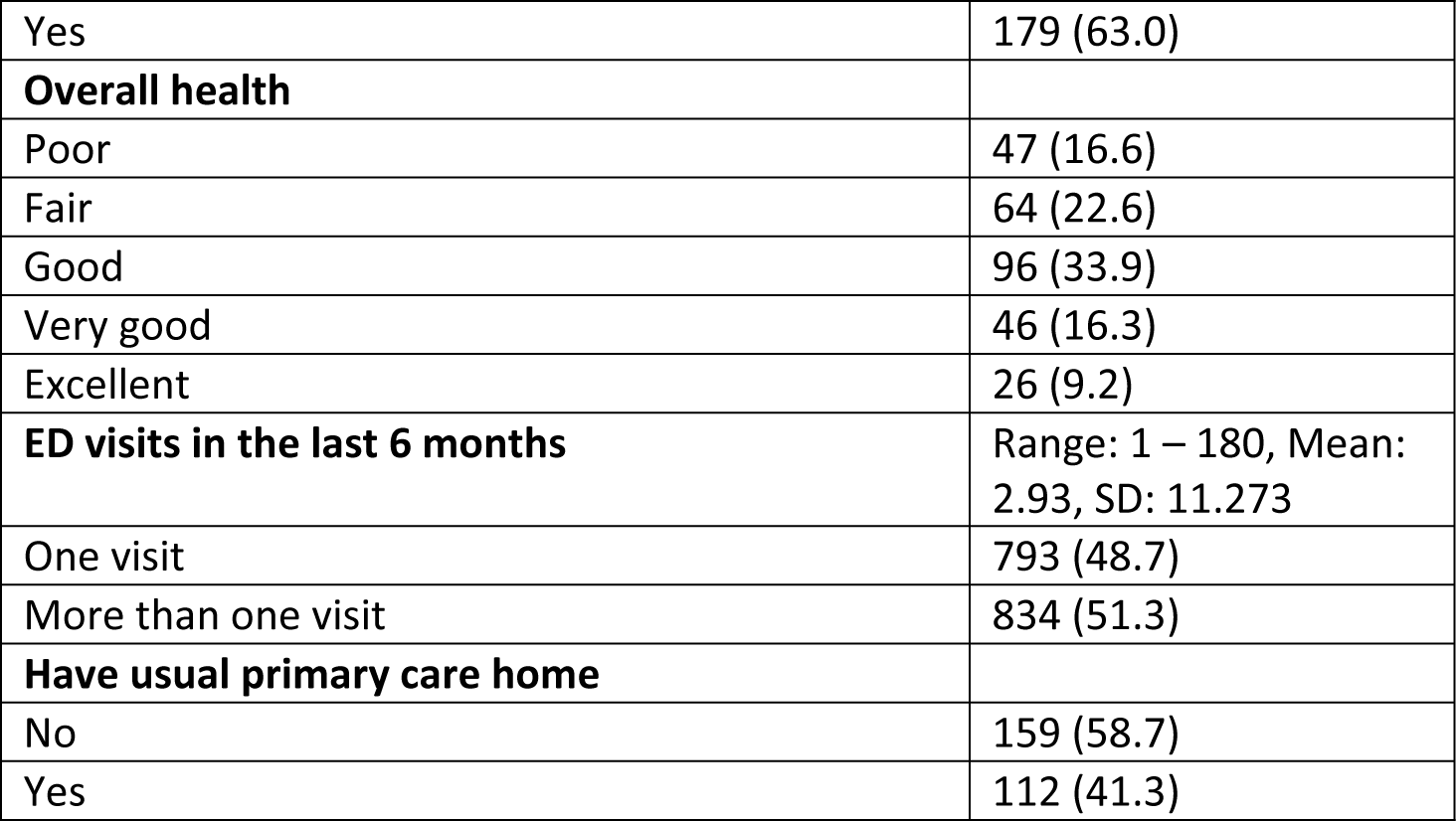
Demographic characteristics of patients completing the EEE-HC Scale (N=284)

The sample collected at SMH included 48.4% of respondents reporting they were not born in Canada, and 42.3% of respondents reporting a first language other than English. In addition, 50.2% of the respondents reported at least somewhat or very difficult financial strain as measured by the Financial Strain Index (Table 4) (58). Consistent with research that indicates that patients in Canadian healthcare settings tend to rate ED care favorably, overall (8, 40, 59), ratings of quality of care in our sample were high, with an average of 8.11 (SD 2.14) on a scale of 0-10, as measured by an item from the British Columbia Emergency Department Patient Experiences of Care scale (8, 21, 31, 60). As reported elsewhere, in an effort to understand experiences of care in a more nuanced way, and in light of deepening health care inequities in Canada (20, 61, 62), the larger *EQUIP Emergency* survey also sought to understand patients’ experiences of discrimination, both in their everyday life and during their ED visit (8, 21, 31). In the study sample, using the Everyday Discrimination Scale (63), 63% reported experiencing some form of discrimination in their everyday lives, however, on a scale of 0 – 45, the mean was relatively low at 8.36 (SD 9.421). Similar to trends seen in the larger study, structurally disadvantaged groups of people reported significantly more discrimination in the SMH ED.

#### Pyschometrics: EEE-HC Scale

The EEE-HC Scale items reflected the key dimensions of EOHC as discussed above. Patients were asked during their visit whether their interactions with health care staff were equity-oriented, using a binary response scale: yes (1) or no (0). The EEE-HC total score is a count of the number of items rated by patients as “yes” (1) for all items except “discriminated against you” which received 1 point in the count for “no” responses, with a range of 0 to 9. Scores on the EEE-HC Scale provide an index of the degree or level of episodic EOHC, from lower to higher.

IRT with a two-parameter (difficulty and discrimination) model was used to examine the item characteristics of the 10-items in the EEE-HC Scale. To test concurrent validity of the EEE-HC Scale, correlations of the total EEE-HC score and quality of care and t-tests of differences in EEE-HC total scores by individual characteristics (age, gender, financial situation, identity as Indigenous or non-Indigenous, and employment status) were conducted. All analyses were conducted in STATA 15.0.

In the IRT analysis of the 10 items of the EEE-HC Scale, one item (“Encourage you to return, if you need to”) had a discrimination score of 0.99. Discrimination scores close to 1.0 indicate that the item does not differentiate between people with varying degrees of the underlying concept. This item’s poor discrimination is likely related to the fact that the item does not apply to all people in the context of care provided in EDs, and so was dropped and the 2-parameter model was re-estimated. Table 5 shows the nine-item EEE-HC Scale, and provides the final model with difficulty, discrimination, and percent of people endorsing each item. As difficulty scores for items decrease, more participants responded “yes” with respect to that item.

**Table 5.** Item difficulty and discrimination parameters from the IRT model and frequency each item was endorsed.

|  | Difficulty | % Responding Yes |
| --- | --- | --- |
| 1. discriminate against you? | -1.79 | 8.8% |
| 2. treat you with courtesy and respect? | -1.44 | 93.3% |
| 3. make you feel welcome? | -1.43 | 89.9% |
| 4. try to make you as comfortable as possible? | -1.09 | 81.9% |
| 5. give you advice that is suitable for you? | -0.96 | 80.7% |
| 6. seem open to talking about what is important to you? | -0.81 | 77.3% |
| 7. learn enough about you to give useful advice? | -0.81 | 76.1% |
| 8. try to help you get services you need? | -0.69 | 68.1% |
| 9. learn about problems you might have getting services (e.g., costs, transportation, getting a referral, etc.)? | 0.45 | 39.1% |

Total scores on the EEE-HC Scale ranged from 0 to 9, with a median of 7.5. Cronbach’s alpha for the 9 items was 0.82. Evidence of concurrent validity, based on a high correlation with quality of care was strong (r=0.61). Table 6 summarizes the differences in EEE-HC scores by participant characteristics. Greater financial strain was associated with lower EEE-HC scores. Those with lower EEE-HC scores were more likely to identify as Indigenous, to have a recent shelter stay, have English as their first language and be unemployed. Scores did not vary by age or gender. The lower scores among people who have English as their first language can be explained, in part, by research showing that newcomers whose first language is not English tend to rate their satisfaction with care quite highly, reflecting appreciation for access to care that may be much less accessible in their countries of origin (64–67). The lower EEE-HC scores experienced by Indigenous peoples are not surprising in the Canadian context, and reflect ongoing research demonstrating the extent to which high proportions of Indigenous people face multiple forms of discrimination and stigma when accessing health care, impacting their experience of care (3, 4, 6, 55, 68).

**Table 6.**
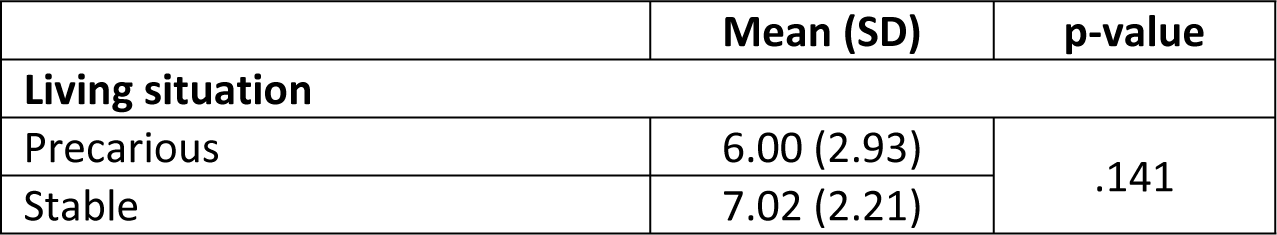
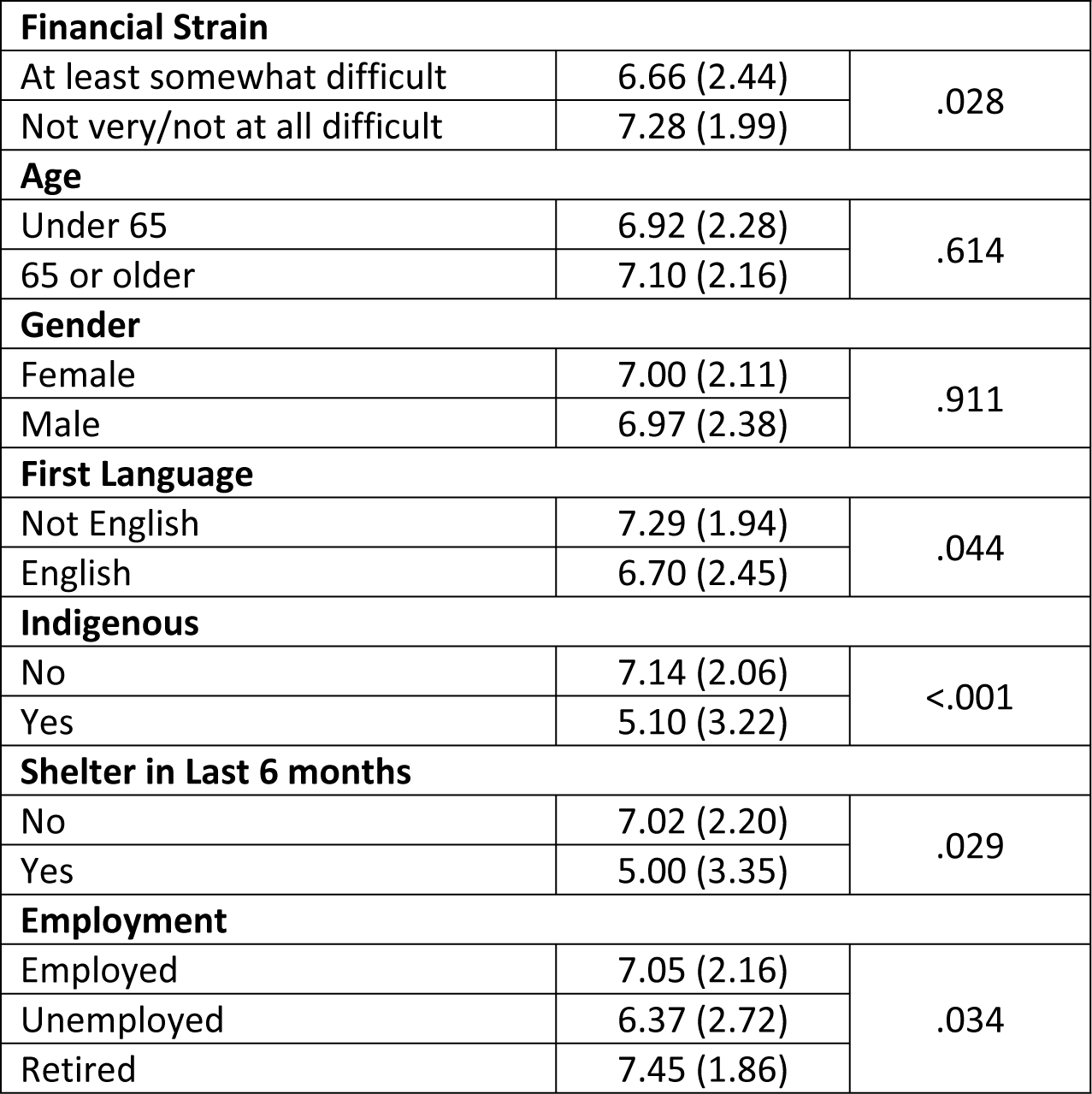
Differences in EEE-HC total score by participant characteristics.

|  | Mean (SD) | p-value |
| --- | --- | --- |
| **Living situation** |  |  |
| Precarious | 6.00 (2.93) | .141 |
| Stable | 7.02 (2.21) |  |
| Financial Strain |  |  |
| At least somewhat difficult | 6.66 (2.44) | .028 |
| Not very/not at all difficult | 7.28 (1.99) |  |
| Age |  |  |
| Under 65 | 6.92 (2.28) | .614 |
| 65 or older | 7.10 (2.16) |  |
| Gender |  |  |
| Female | 7.00 (2.11) | .911 |
| Male | 6.97 (2.38) |  |
| First Language |  |  |
| Not English | 7.29 (1.94) | .044 |
| English | 6.70 (2.45) |  |
| Indigenous |  |  |
| No | 7.14 (2.06) | <.001 |
| Yes | 5.10 (3.22) |  |
| Shelter in Last 6 months |  |  |
| No | 7.02 (2.20) | .029 |
| Yes | 5.00 (3.35) |  |
| Employment |  |  |
| Employed | 7.05 (2.16) | .034 |
| Unemployed | 6.37 (2.72) |  |
| Retired | 7.45 (1.86) |  |

Respondents with a first language other than English had significantly higher EEE-HC scores compared to those who had English as their first language spoken (Table 6). The context of SMH is such that it serves a high proportion of newcomers and people with a mother tongue other than English (49.4%), relative to other regions in the province (69). Research shows that once newcomers are able to access care, they tend to rate their care received in Canada highly and report high levels of trust in their providers (64–67). Thus, this pattern of higher EEE-HC scores among people with a first language other than English aligns with published literature illustrating overall high ratings of care among newcomer groups, for many of whom English is not their first language (55, 69).

## Discussion & implications for both scales

We have systematically used an equity lens to study people’s experiences of care, and from that identified the key dimensions of EOHC, and derived items to measure experiences that align with those key dimensions (9, 14). Consequently, we have developed two scales, one for use in settings where the care relationship is ongoing, such as primary care, and another for use in settings where care is episodic, such as EDs or walk-in clinics.

There continue to be strong calls to integrate attention to equity in health care provision, and emerging evidence of positive impacts for patients, providers and organizations. However, without access to brief and reliable ways of measuring whether equity-enhancing innovations have intended impacts, and for whom, organizations will continue to face significant challenges justifying and funding such initiatives. Both the EHoCs and EEE-HC Scale are brief, easy-to-administer patient self-report scales that can support organizations to effectively monitor quality of care using an intersectional equity lens. Embedding such scales in CQI processes, and tracking responses over time, may support, shift or expand the ways in which quality of care is conceptualized, enhanced, defined and measured. For example, in a recent CQI initiative at a primary care clinic serving women experiencing major social disadvantages and marginalization, items from the EHoCs were used to tap into women’s perspectives regarding the quality of care received (70). The analysis of women’s responses was particularly useful in highlighting those aspects of care that women rated most highly (e.g., promoting emotional safety and trust), and helping clinicians identify domains of EOHC they seemed to excel in supporting, and those aspects of care needing further improvement. While the EHoCS was developed for use in PHC care settings, it may be appropriate in settings where care is provided by individual providers, for example, physicians or nurse practitioners, where the goal is to provide care over time with a roster of patients.

Given that the items in both scales focus on experiences of care in varied settings, we have also explored the use of these measures beyond health care services. For example, in a study assessing the addition of TVIC as a key dimension of EOHC in educational contexts, we added relevant items from the EHoCs to the Attitudes Related to Trauma Informed Care Scale (ARTIC), a pre-existing scale assessing trauma-informed practice (71, 72). Indeed, as found in a recent scoping review (73), a key limitation of measures to assess EOHC concepts, such as TVIC, is that they rarely include items focused on stigma, racism and discrimination as structural or systemic influences on care experiences. We therefore encourage the testing and use of the EHoCS and the EEE-HC Scale as measures of EOHC beyond health care settings.

In view of ongoing claw-backs in Canadian primary care sectors (and in other international jurisdictions), diminishing opportunities for care based on a continuing relationship with a primary care provider or agency, and increasing shifts toward episodic health care delivery, the availability of both scales makes it possible to measure EOHC in a wide range of contexts. These ways of describing PEOC will be important for research, for quality improvement and monitoring, and potentially, for informing ongoing health care reforms. Both scales have the capacity to shed light on experiences of care using an intersectional lens – providing a more nuanced understanding of EOHC -- versus focusing on a single dimension. Use of these scales can be helpful in highlighting how peoples’ intersecting social locations impact their experiences of care. For example, the EEE-HC Scale allowed us to examine those aspects of care that people valued most highly in EDs (8). Bringing a health equity lens to analyses of PEOC is especially important to inform strategies and recommendations to enhance care. Further testing will enable ongoing refinements to both scales, and provide important insights regarding their acceptability, feasibility, reliability and validation in diverse settings.

## Data Availability

The datasets generated and/or analyzed during the current study are not publicly available because we are still actively working on analyses, but are available from the corresponding author on reasonable request.

## Acknowledgements

We extend our gratitude to the many patients, staff and organizations with whom we have collaborated on research in the primary health care and emergency department sectors. Thank you to Cheyanne Stones for working so ably with our research teams. Nadine Wathen was supported by a SSHRC (Tier 1) Canada Research Chair.

